# Estimating the undetected burden of infections and the likelihood of strain persistence of drug-resistant *Neisseria gonorrhoeae*

**DOI:** 10.1101/2024.08.28.24312729

**Authors:** Kirstin I. Oliveira Roster, Minttu M. Rönn, Heather Elder, Thomas L. Gift, Kathleen A. Roosevelt, Joshua A. Salomon, Katherine K. Hsu, Yonatan H. Grad

## Abstract

Antimicrobial resistance (AMR) is a serious public health threat. *Neisseria gonorrhoeae* has developed resistance to all antibiotics recommended for treatment and reports of reduced susceptibility to ceftriaxone, the last-line treatment, are increasing. Since many asymptomatic infections remain undiagnosed and most diagnosed infections do not undergo antibiotic susceptibility testing, surveillance systems may underestimate the number of ceftriaxone non-susceptible infections. There is an urgent need for better interpretation and use of surveillance data to estimate the prevalence of resistance. In this modeling study, we simulated the spread of a new strain of ceftriaxone non-susceptible gonorrhea in a population of men who have sex with men as well as heterosexual men and women. We compared scenarios with varying strain characteristics and surveillance capacity. For each scenario, we estimated (i) the number of undetected infections by the time the non-susceptible strain was discovered and (ii) the likelihood of strain persistence in the absence of newly reported cases. Upon detection of one ceftriaxone non-susceptible isolate, the undetected disease burden was an estimated 5.4 infections with substantial uncertainty (0-18 infections, 95% uncertainty interval). In the absence of additional reports of ceftriaxone non-susceptible infections over the subsequent 180 days, the estimate declined to 2.5 infections with a narrower uncertainty interval (0-10 infections). The likelihood of ongoing transmission also declined from 66% (26-86%) at first detection to 2% (0-10%) after 180 days. Enhanced monitoring of the magnitude and trends of AMR is a key priority to reduce the burden of resistance and extend the useful lifespan of last-line antibiotics.

## Introduction

The urgent public health threat of antimicrobial resistance (AMR) demands new strategies to slow and control its spread. An estimated 1.27 million people died from resistant bacterial infections in 2019 globally (1). Especially concerning are rising rates of resistance to last-line antibiotics (2) and resistance in pathogens with high incidence, such as *Mycobacterium tuberculosis, Salmonella*, and *Neisseria gonorrhoeae* (3). *N. gonorrhoeae* causes an estimated 82 million infections annually (4) and has developed resistance to all antibiotics recommended for treatment (5). While novel therapeutics are in development (6,7), reports of resistance to ceftriaxone, the last approved treatment, are increasing (8–12), including in the United States (13).

Two cases of a *N. gonorrhoeae* strain with reduced susceptibility to ceftriaxone and resistance or reduced susceptibility to all other approved anti-gonococcal antibiotics were identified in Massachusetts in 2022 (14). After serendipitous discovery of the first case through urine culture, the Massachusetts Department of Public Health (MDPH) collaborated with the originating microbiology laboratory and the US Centers for Disease Control and Prevention (CDC) to perform retrospective molecular testing of remnant specimens from nucleic acid amplification tests (NAATs) conducted between January and September 2022; a total of 54 remnant specimens were tested, leading to the discovery of a second case from the same community health center from which the first non-susceptible case was identified. The MDPH also issued a clinical alert (January 2023) to encourage increased gonococcal culture for symptomatic patients across Massachusetts (14,15). The expanded culture surveillance has yet to identify any additional ceftriaxone non-susceptible cases.

However, many asymptomatic infections are undiagnosed (16–18) and the vast majority of diagnosed gonococcal infections do not undergo antibiotic susceptibility testing (19). Even after the clinical alert, culture isolates associated with only 5.5% of all NAAT-positive gonococcal cases in Massachusetts were submitted to the MDPH (540 specimens from 9859 cases in the first year following clinical alert release) (20). Consequently, surveillance systems may underestimate the number of ceftriaxone non-susceptible cases. There is an urgent need for better interpretation and use of surveillance data to estimate the unseen prevalence of resistance.

In this modeling study, we sought to understand how surveillance signals relate to the true infection burden, including how much time must pass without detecting new cases of the novel strain before being confident that there is no ongoing cryptic transmission. To do so, we developed a mathematical model of gonorrhea transmission in Massachusetts among men who have sex with men (MSM), men who have sex with men and women (MSMW), men who have sex with women (MSW), and women who have sex with men (WSM). We further split each group into two sexual activity classes with either high or low partner change rates and calibrated mixing parameters to MDPH surveillance data. The model accounted for the natural history of gonococcal infection, care seeking behavior, and surveillance of antimicrobial resistance. We simulated the introduction of a ceftriaxone non-susceptible case and measured the number of unreported infections and the likelihood of cryptic transmission for different surveillance scenarios, including the observed scenario in Massachusetts, in which no cases have been found since detection of two ceftriaxone non-susceptible infections in 2022. As sensitivity analyses, we examined scenarios in which we varied treatment failure rates of the non-susceptible strain and rates of asymptomatic screening and antibiotic susceptibility testing.

This study provides a roadmap for situations where increased resistance to the last-line antibiotic is observed globally and surveillance systems are tasked with tracking early introductions and emergence of resistance locally. Improved interpretation of gonorrhea surveillance data can help guide surveillance systems of AMR more broadly. Monitoring population prevalence of antibiotic resistance is fundamental to ensure treatment guidelines are compatible with the resistance profiles of circulating strains, to respond quickly to rising levels of resistance, and to extend the clinically useful lifespan of last-line antibiotics (3,19,21–23).

## Methods

### Transmission model

We developed a stochastic susceptible-infected-susceptible compartmental model of gonorrhea transmission in Massachusetts (**Figure S1**). We accounted for symptomatic and asymptomatic infections with either a ceftriaxone-susceptible or non-susceptible strain of gonorrhea. In the context of this model, we defined strains in terms of their antibiotic susceptibility patterns, distinguishing specifically between those susceptible and non-susceptible to treatment with 500mg ceftriaxone. We assumed that there were no infections with multiple strains of *N. gonorrhoeae*. The population was stratified into eight subgroups based on gender identity, gender identity of sex partners, and two sexual activity groups with either high or low partner change rates.

Gonococcal infections were diagnosed in response to symptomatic care seeking or asymptomatic screening. Resistance was monitored through antibiotic susceptibility testing of a subset of gonorrhea-positive isolates, whereby not only the infection but also the strain causing the infection was identified by the surveillance system. Rates of asymptomatic screening (by gender, gender of sex partners, and sexual activity group) and antibiotic susceptibility testing (by gender) were among the parameters fitted to surveillance data in the main analysis (**Table S2**). In sensitivity analyses, we evaluated i) rates of antibiotic susceptibility testing ranging from 1-40% of detected cases and ii) screening frequency with up to two-fold increases (preserving the relative screening intensity between groups).

We assumed that treatment of ceftriaxone non-susceptible infections results in treatment failure in 80% of infections. In sensitivity analyses, we considered treatment failure rates of 30-90%. This partial treatment failure is concordant with the two cases detected in Massachusetts, since these were successfully treated with ceftriaxone, the first case with 500mg and the second case with 1g plus 1.2g azithromycin (14). We assumed that identification of treatment failure (either through persistent symptoms or test of cure of asymptomatic infections) would trigger antibiotic susceptibility testing, leading to the detection of the non-susceptible infection, and treatment with an alternative curative antibiotic. Asymptomatic cases that did were not followed up with a test of cure remained infectious until natural recovery. Treatment of infections with the susceptible strain was always successful.

### Model Calibration

The model was calibrated to gonorrhea diagnosis rates by gender and prevalence among MSM, MSW, and WSM using an Approximate Bayesian Computation (ABC) rejection sampling approach with 1 million iterations (see **Table S1** for starting conditions and **Table S3** for calibration targets). Prior distributions of model parameters were defined using surveillance data from MDPH, where available, and values from the literature otherwise (see **Table S2** for prior and posterior parameter distributions of fitted parameters and **Table S1** for fixed parameters). Calibration was performed for the ceftriaxone-susceptible strain only.

### Introduction of non-susceptible strain

After calibration of the ceftriaxone-susceptible strain dynamics, we introduced a single index case of the non-susceptible strain into one of the eight population subgroups and ran 500 simulation iterations for each subgroup. Sensitivity analyses were performed on the calibrated model. We iteratively filtered simulations that were consistent with surveillance scenarios where detection of the first case was followed by increasing periods (1-250 days) without reports of ceftriaxone-non-susceptible infections. We examined scenarios where more than 1 case was detected initially before detection ceased, from 2 detected cases, as was observed in Massachusetts, up to 10 detected cases.

For each scenario, we calculated (1) the cumulative number of undetected infections with the non-susceptible strain and (2) the likelihood of elimination of the non-susceptible strain, defined as the proportion of simulations without new undetected infections.

Analyses were performed using python 3.9.13. All code is available at https://github.com/gradlab/iceberg.

## Results

### First discovery of the non-susceptible strain

A diversity of transmission scenarios yielded similar surveillance signals (**Figure 1**). Three example simulations (**Figure 1**) showcase the wide range of possible trajectories underlying the detection of 2 cases of the ceftriaxone non-susceptible strain, followed by a period without new reports of non-susceptible infections. The strain may be eliminated shortly after detection of the second case (**Figure 1A**) or it may persist in the population via continued transmission (**Figure 1B**). Detection of additional cases was associated either with strain persistence (**Figure 1B**) or strain elimination (**Figure 1C**).

**Figure 1.**
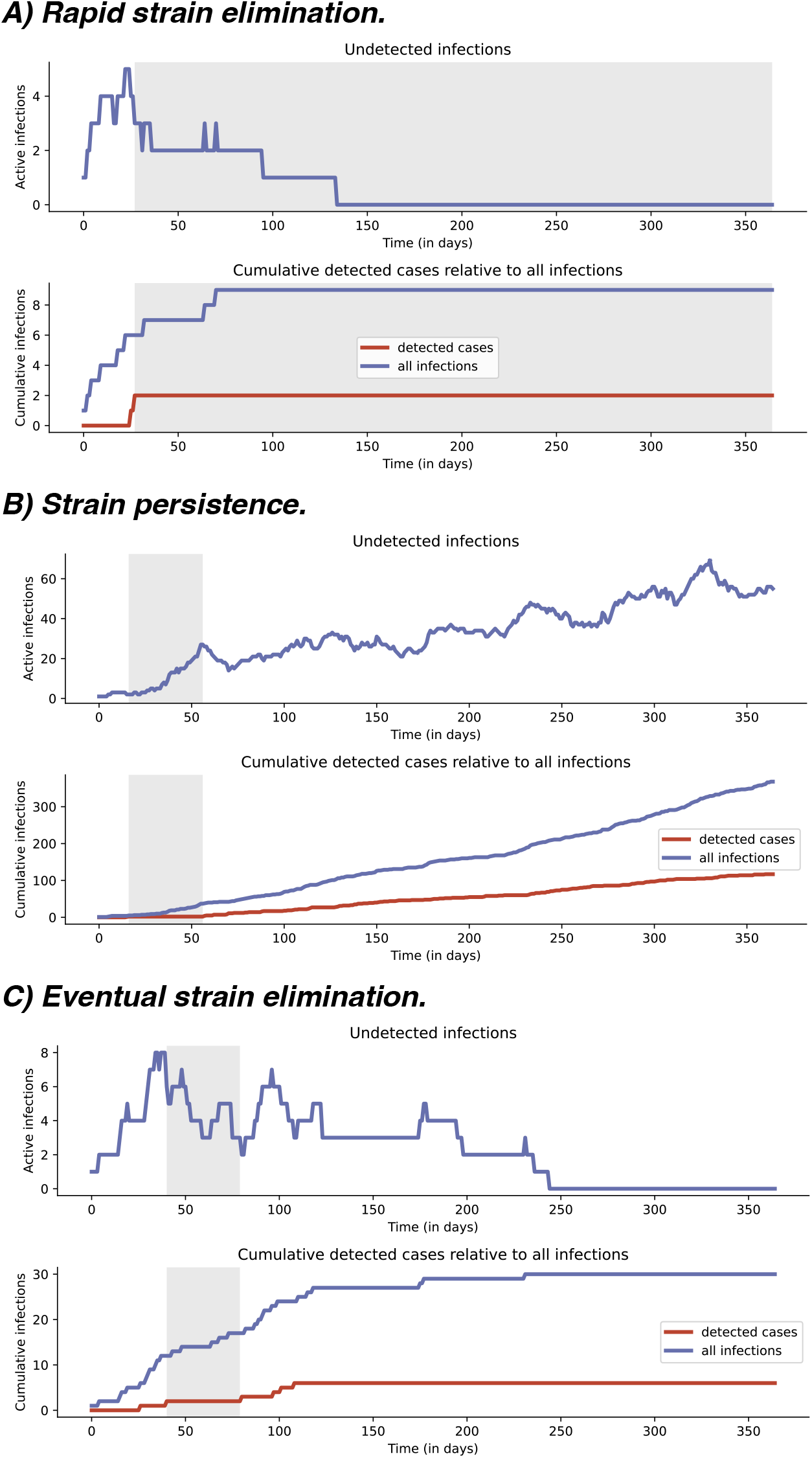
Sample simulation trajectories illustrating the relationship between observed and hidden dynamics. Active undetected infections (top row of the panel) and cumulative detected and total infections (bottom row of the panel) of the non-susceptible strain over time for three sample simulations. Shaded areas (gray) indicate time periods in which exactly two non-susceptible cases have been detected.

The range of possible trajectories led to wide uncertainty intervals around initial estimates of the number of unreported infections. Upon detection of the first non-susceptible case, the total number of undetected non-susceptible infections was estimated to be 5.4 infections with a 95% uncertainty interval ranging from 0 to 18 infections (**Figure 2**). Over time, additional detections of the new strain or the absence of new detections narrowed the range of possible trajectories and thus increased confidence in estimates of the number of undetected infections. In our simulations, after 6 months (180 days) without any new detections of non-susceptible infections, the estimate of the cumulative number of unreported infections was reduced to 2.5 with a narrower 95% uncertainty interval of 0-10 infections (**Figure 2**).

**Figure 2.**
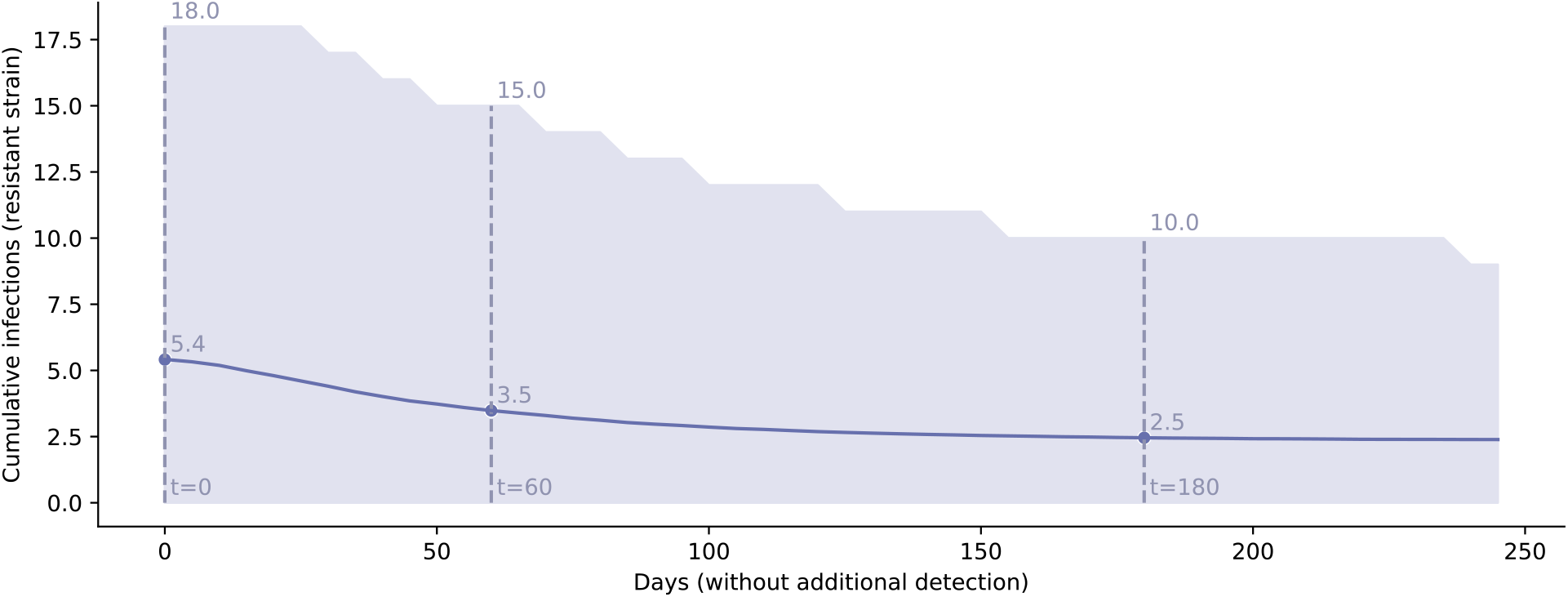
Updating the estimate of the undetected burden of non-susceptible infections with the passing of time without new reported non-susceptible infections after detection of one case of ceftriaxone non-susceptible strain. Estimate of the cumulative number of undetected non-susceptible infections as the number of days without observed non-susceptible cases increases. The solid line represents the average estimate and the shaded area highlights 95% uncertainty interval. Dashed lines highlight values at 0, 60, and 180 days without detection.

Increasing time without new detections of non-susceptible infections also raised the likelihood that the strain had been eliminated. On the day the first non-susceptible case was detected, the likelihood of elimination was only 34% (14-74%, 95% uncertainty interval), meaning that the new strain continued causing new infections in 66% of simulations (**Figure 3**). Each additional day without detection reduced the likelihood that the non-susceptible strain still circulated in the population. After 180 days without detection of ceftriaxone non-susceptible cases, the strain had been eliminated in 98% of simulations (90-100%, 95% uncertainty interval).

**Figure 3.**
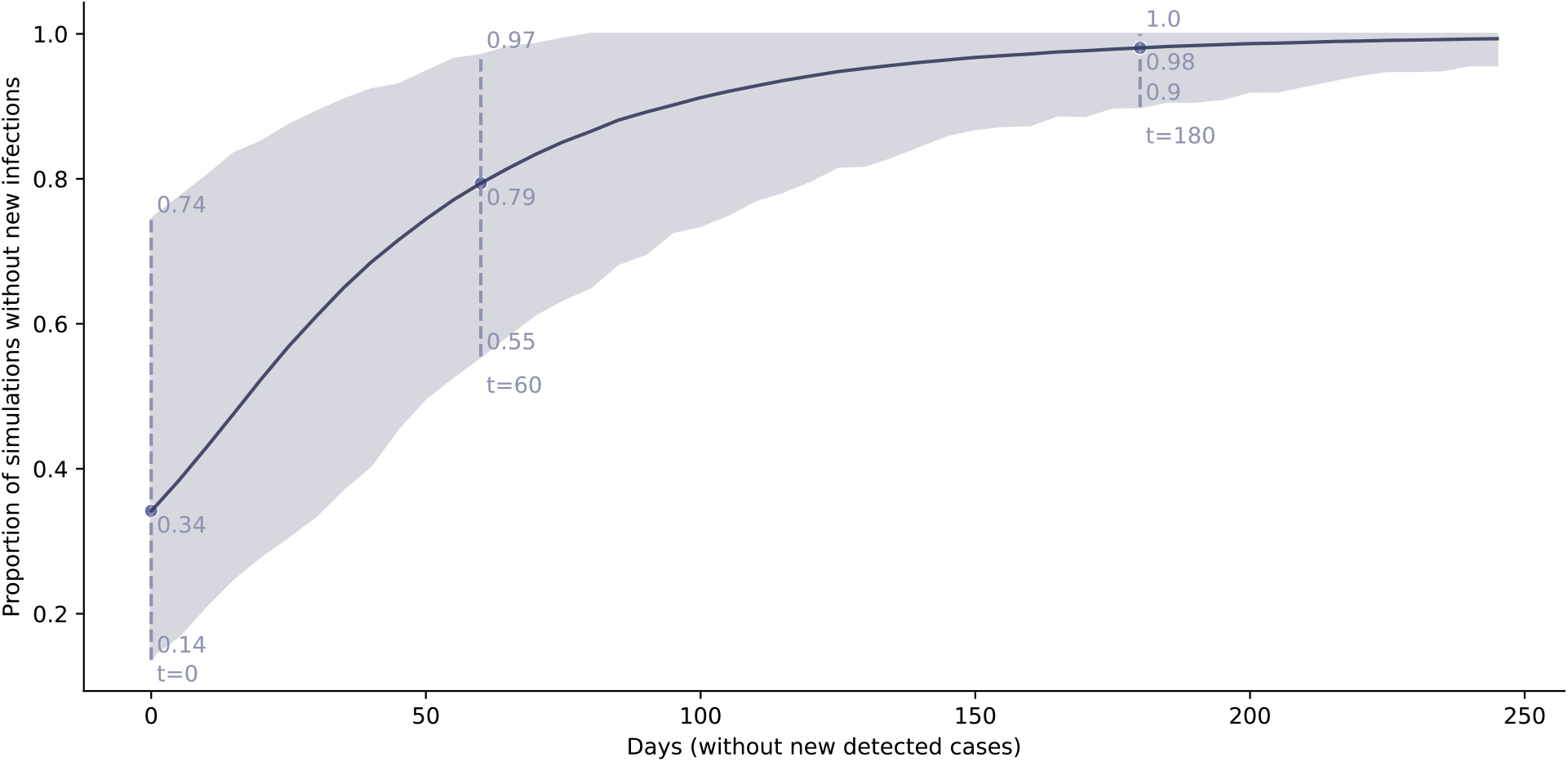
Likelihood of strain elimination for increasing days without detection of ceftriaxone non-susceptible cases after detection of one case of ceftriaxone non-susceptible strain. Proportion of simulations without new undetected infections for increasing days on which no new cases were detected (mean and 95% uncertainty interval). Dashed lines highlight values at 0, 60, and 180 days without detection.

### Massachusetts scenario: detecting 2 non-susceptible infections

Considering the scenario observed in Massachusetts in 2022, the number of unreported infections upon detection of 2 non-susceptible infections was estimated to be 10.6 infections (2-28, 95% uncertainty interval) (**Figure S2a**). In the absence of additional detections of ceftriaxone non-susceptible infections over the subsequent 180 days, the estimate declined to 5.7 cumulative undetected infections (1-16, 95% uncertainty interval). The likelihood of strain elimination increased as no new cases were detected, from 20% (7-53%, 95% uncertainty interval) on day 0 to 98% (88-100%, 95% uncertainty interval) on day 180 (**Figure S2b**).

### Comparing scenarios with varying numbers of initially detected cases

Expanding upon the scenarios with 1 and 2 detections, we examined whether the impact of time without detection depends on the number of cases initially detected. We considered simulations with initial detections ranging from 1 to 10 cases, followed by periods without new reports of non-susceptible infections, placing no restrictions on the time frame in which these initial infections occurred. The cumulative number of infections varied primarily by the number of initial detections (**Figure 4a**), whereas the average likelihood of strain elimination varied primarily by the time since the last detected case (**Figure 4b**). When 1 case was discovered, it took 100 days to be 90% confident that the non-susceptible strain had been eliminated, whereas when 10 cases had been reported, it took 150 days to achieve 90% elimination likelihood (**Figure 4b**). Long time periods without new detected cases became increasingly unlikely as more initial cases were detected, resulting in wider uncertainty intervals around estimates of the undetected infection burden and the likelihood of strain elimination (**Figure S3**).

**Figure 4.**
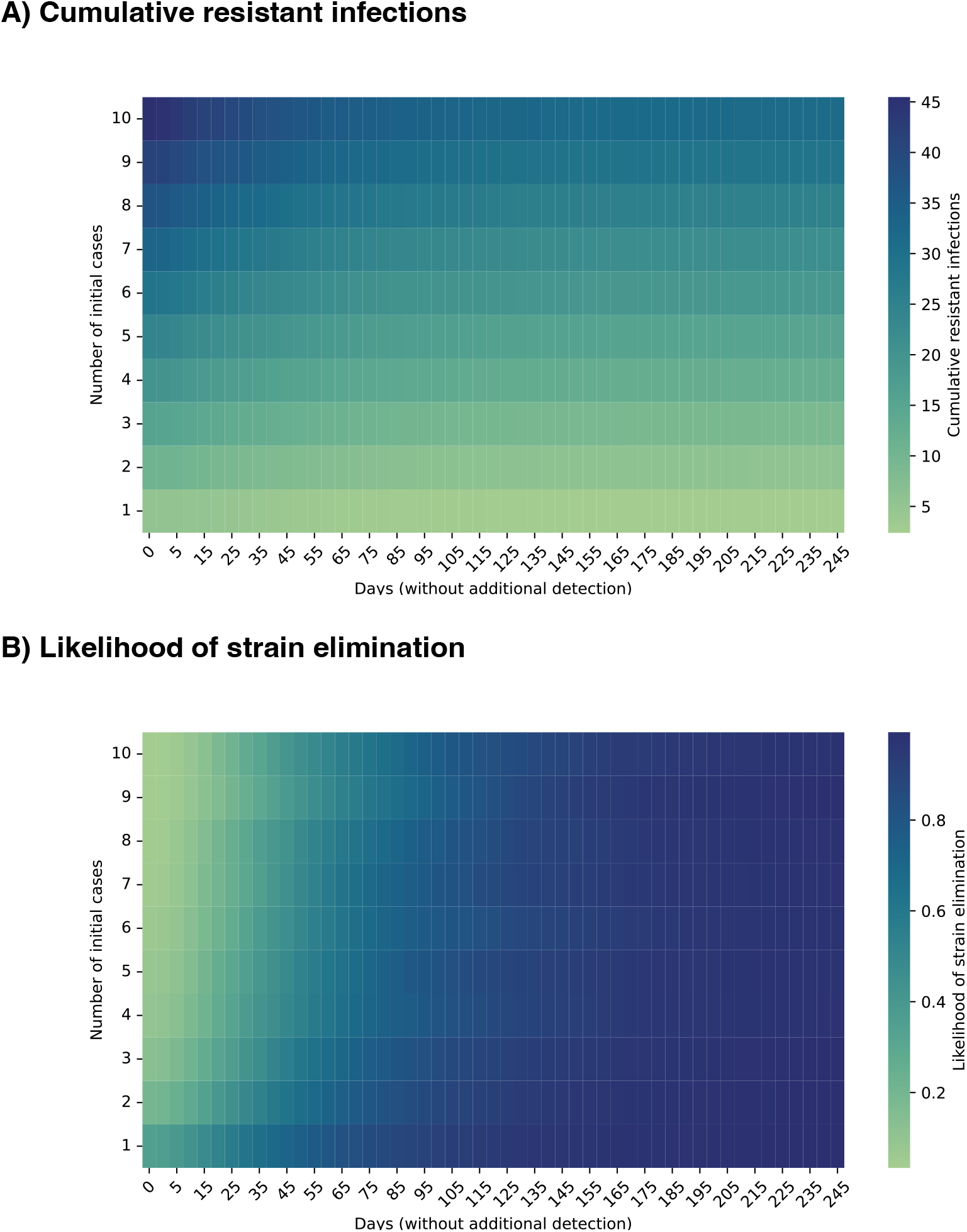
Varying numbers of initial detections followed by increasing periods without new detected cases. Estimates of the average (A) cumulative number of infections and (B) likelihood of strain elimination for scenarios with 1-10 initial detected cases followed by 0-245 days without detection of new cases.

### Comparing strains with varying levels of resistance

Non-susceptible strains were detected if a specimen was randomly sampled for antibiotic susceptibility testing or through treatment failure. When a strain had a high treatment failure rate, infections were less likely to be successfully treated and therefore more likely to be detected, due to either persistent symptoms or test of cure. Higher treatment failure rates were therefore associated with a lower number of unreported infections (**Figure S4**). Conversely, not detecting any new cases of strains with a high treatment failure rate was more likely a sign that there were indeed no new infections, resulting in higher confidence in strain elimination. After 30 days without detecting new cases, a strain with a treatment failure rate of 90% had been eliminated in 62% of simulations (36-90%, 95% uncertainty interval), whereas a strain with a treatment failure rate of 30% had only a 47% chance of being eliminated (24-79%, 95% uncertainty interval) (**Figure 5**).

**Figure 5.**
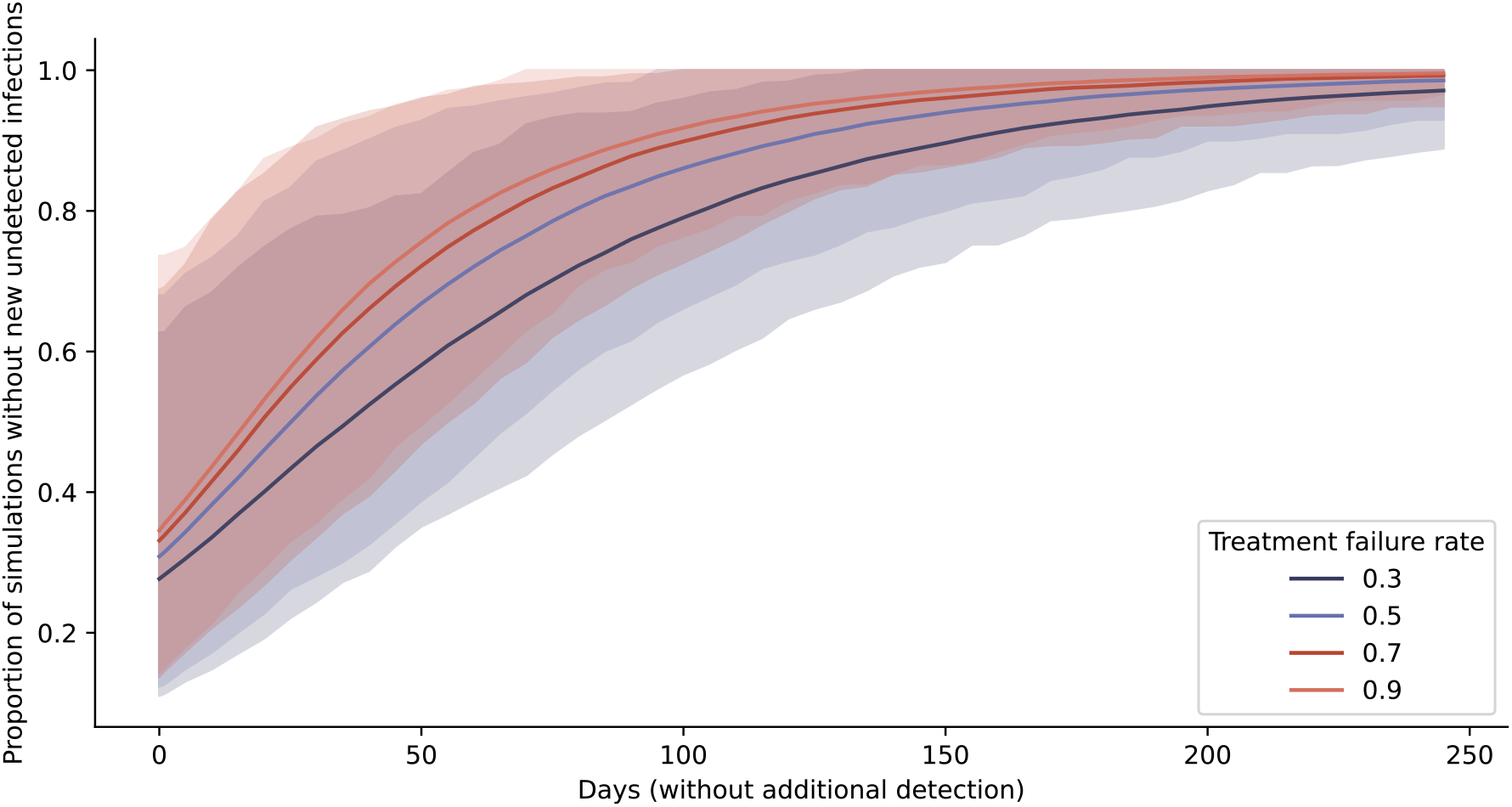
Varying treatment failure rates of non-susceptible strain. Likelihood of strain elimination for increasing days without newly reported non-susceptible cases (mean and 95% uncertainty interval), for treatment failure rates of 30%, 50%, 70%, and 90%.

### Comparing varying surveillance intensities

We examined to what extent our estimates of ceftriaxone non-susceptible infections depended on antibiotic susceptibility testing coverage and screening frequency. Increasing levels of surveillance via increased antibiotic susceptibility testing and asymptomatic screening both reduced the estimated number of undetected infections, though the effect of increasing screening was stronger (**Figure S5**). Doubling the baseline screening intensity (with fixed antibiotic susceptibility testing of 10%) lowered the estimated undetected burden on the day of the first detection from 5.1 (0-17, 95% uncertainty interval) to 4.3 infections (0-15, 95% uncertainty interval), whereas doubling the rate of antibiotic susceptibility testing lowered the estimated undetected infections to only 4.8 (0-16, 95% uncertainty interval) (**Figure S5**). Increased surveillance also reduced the likelihood of ongoing transmission in the absence of detection, again with a stronger effect of asymptomatic screening over antibiotic susceptibility testing. Doubling screening frequency raised the likelihood of strain elimination from 34% (17-66%) to 41% (21-77%), while doubling the antibiotic susceptibility testing rate raised the likelihood to 35% (17-69%) (**Figure 6**).

**Figure 6.**
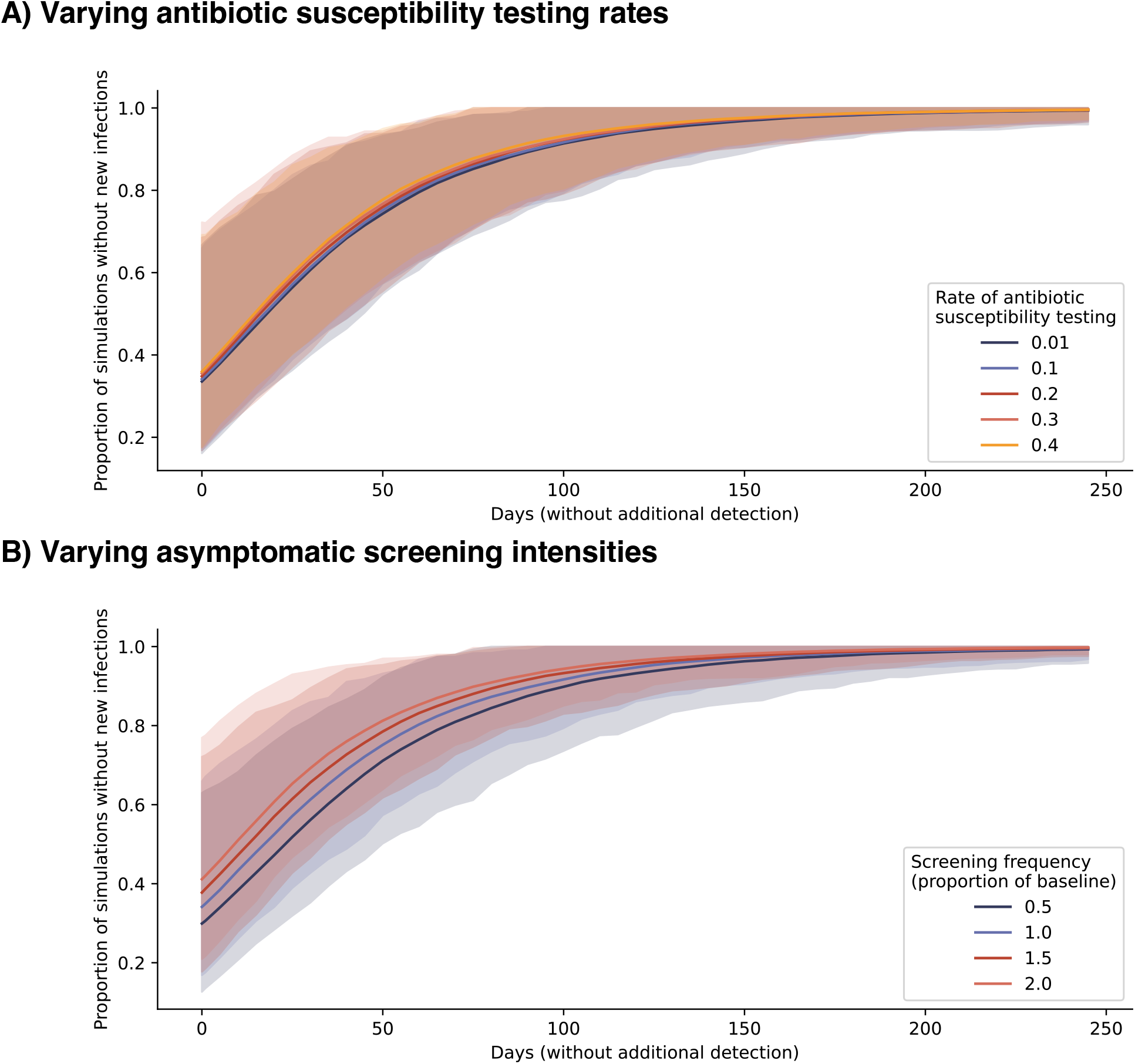
Varying surveillance intensities. Likelihood of strain elimination for increasing days without newly reported non-susceptible cases (mean and 95% uncertainty interval), for (A) antibiotic susceptibility testing rates ranging from 1-40% of all positive gonorrhea diagnoses, and (B) asymptomatic screening rates ranging from 50-200% of baseline levels in the calibrated model, preserving the relative screening intensities by population group.

## Discussion

To help guide the surveillance of AMR, we estimated how many infections evade detection when a new ceftriaxone non-susceptible strain of *N. gonorrhoeae* is first detected, taking the reports of two multidrug non-susceptible infections in Massachusetts as a case study. In our simulations, a wide range of disease scenarios generated similar surveillance signals that were consistent with 2 detected cases followed by a period without detection; these hidden dynamics ranged from rapid strain elimination to persistence of the strain in the population. This diversity of outbreak trajectories compatible with the same surveillance signal illustrates how difficult it can be to interpret both days with and without detection and to anticipate whether newly detected strains of concern will continue to spread in a given location.

We estimated that on the day a ceftriaxone non-susceptible strain was first discovered in Massachusetts, the undetected burden was 5.4 infections and in the range of 0 to 18 cases. By the time two infections had been detected, an estimated 10.6 infections had evaded detection, with a likely range of 2-28 infections. Transmission was ongoing in 66% of simulations the day the first case was detected and declined to 2% after 180 days (six months) without new reports of non-susceptible cases. In simulations with two detected infections, 80% of simulations had ongoing transmission the day the second case was detected, and the likelihood of strain persistence also declined to 2% after 180 days without new reports of ceftriaxone non-susceptible infections.

The link between surveillance signals and the true disease dynamics was stronger for highly resistant strains. When strains had a higher risk of treatment failure, the absence of detection was more indicative of an underlying absence of transmission than for intermediately resistant strains. This finding suggests that the phenotypic characteristics of the novel strain should be considered in the interpretation of surveillance data.

The marginal improvement in confidence of elimination for each day without detecting a new case decreased as more non-susceptible cases were reported. When only 1 case had been detected, it took 100 days to be 90% confident that the strain had been eliminated. But if 10 cases had initially been reported, the likelihood of strain elimination after 100 days without detection was only 76%, and it would take 150 days without detection to reach 90% confidence. This finding emphasizes the importance of responding quickly to the first detected cases of a new strain of concern.

Increasing asymptomatic screening frequency reduced the estimated undetected infection burden and thus increased surveillance accuracy. Greater screening coverage also reduced the likelihood of ongoing transmission in the absence of detection, likely due to the longer infectious period of asymptomatic infections which can produce new transmission events even after a period without new detections of non-susceptible infections.

Our findings rely on several modeling assumptions. We maintained many of the structural elements in prior gonorrhea models (24–27) and added complexity only as necessary. Many models of gonorrhea focus on a single subpopulation, such as men who have sex with men (24,28). We modeled both heterosexual and homosexual contact networks jointly, since ceftriaxone resistance has been observed in heterosexual men and women (8), while gonorrhea prevalence is highest among MSM (29). However, we did not explicitly account for other population heterogeneities such as age structure, geography, or infection prevalence by anatomic site, since we did not have sufficiently confident estimates of parameters required to construct such a model.

The rate at which gonorrhea infections are imported to Massachusetts (and globally) is poorly understood. In this model, we measured outcomes for a single importation event, a realistic assumption if importations of non-susceptible strains are sufficiently infrequent. Estimates of the cumulative burden and likelihood of strain elimination will differ if multiple independent importations occur or if non-susceptible infections are replenished before the strain is eliminated. Given that reduced susceptibility to ceftriaxone has been reported only 3 times in the US to date (13,14), it is probable that the two infections observed in Massachusetts were linked. We assumed that importation may occur in any of our 8 subpopulations, since the relative contribution of different genders, genders of sex partners, and sexual activity groups to strain importation has not yet been characterized. Once more data become available, the simulations should be reweighted to account for differences in importation frequency. We also note that while focused here on importations of ceftriaxone non-susceptible strains, our results would also apply to *de novo* mutations originating within the population (5).

With increasing global reports of non-susceptibility to ceftriaxone, the last remaining antibiotic for empiric treatment of gonorrhea, challenges in treating gonococcal infections are a serious public health threat. Scaling up surveillance of antimicrobial resistance in gonorrhea is costly, given the challenges with culture-based surveillance, asymptomatic infections, and contact tracing. To optimize the use of limited resources, we need tools to help estimate the infection landscapes that could underlie the data from our surveillance systems. The inferential approach reported here helps fill this need, both in interpreting the data from Massachusetts and in establishing a platform for other settings.

## Data Availability

Code and data are available at https://github.com/gradlab/iceberg.

## Acknowledgments

This project was funded by the U.S. Centers for Disease Control and Prevention, National Center for HIV/AIDS, Viral Hepatitis, STD, and TB Prevention Epidemiologic and Economic Modeling Agreement (NEEMA, #5 NU38PS004651). The findings and conclusions in this report are those of the authors and do not necessarily represent the official position of the CDC or other authors’ affiliated institutions.

