## supplementary_materials for "Estimating the undetected burden of infections and the likelihood of strain persistence of drug-resistant *Neisseria gonorrhoeae*"

### Supplemental Materials and Methods

#### Figure S1. Compartmental model structure

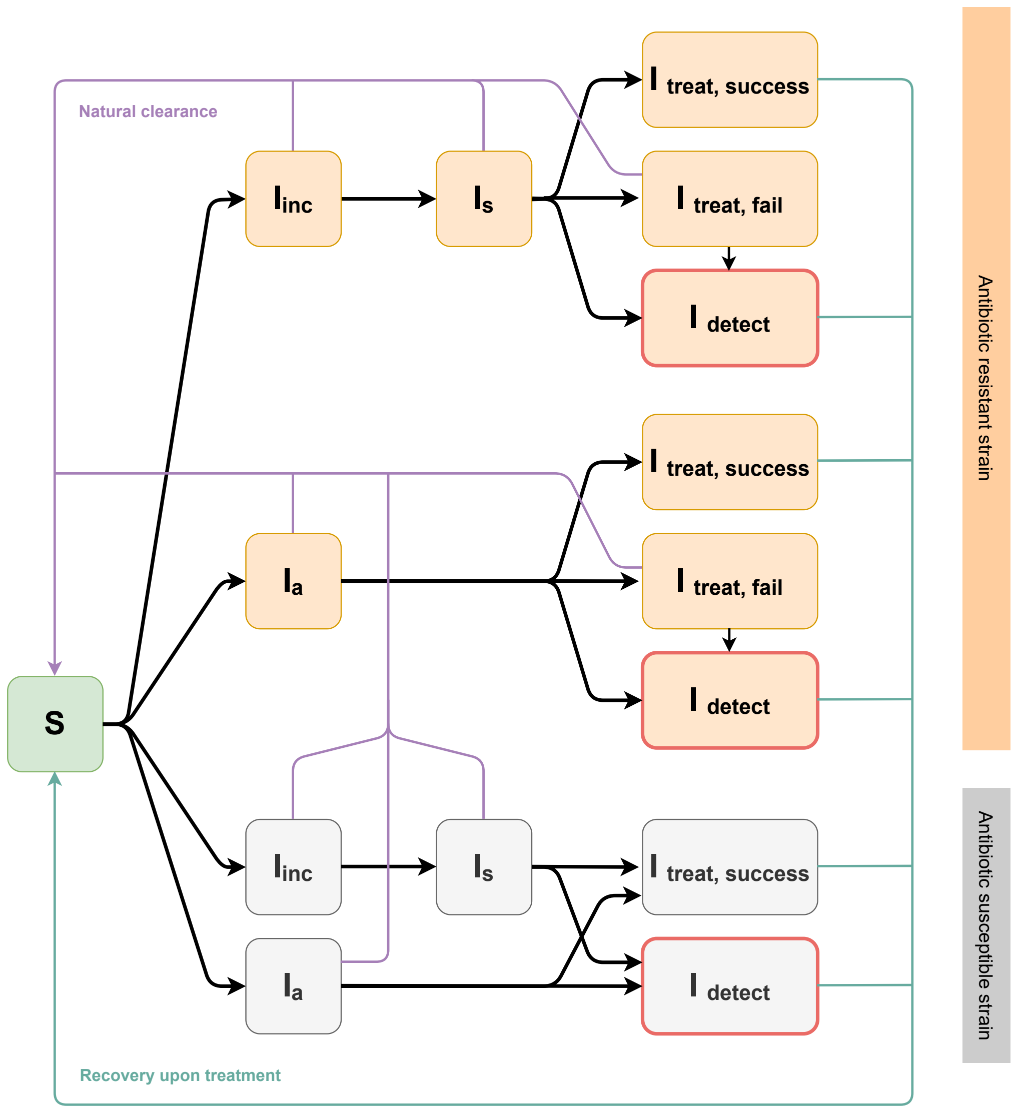

#### Model equations

Susceptible

$\frac{dS}{dt}$ = + population entry - population exit – infection + innate recovery + recovery upon treatment + retreatment

$$\frac{dS}{dt} = \mu_{entry}N - \mu_{exit}S - \frac{\beta IS}{N} + \frac{1}{D_n}(I_{inc} + I_a + I_s + I_{treat,fail}) + \frac{1}{D_r}I_{treat,success} + p_{screen}\, I_{a,r,treat,fail}$$

Symptomatic infection with resistant strain:

$\frac{dI_{inc,r}}{dt}$ = - population exit + infection – innate recovery – symptom onset

$$\frac{dI_{inc,r}}{dt} = p_{symptoms}\frac{\beta S I_r}{N} - \frac{1}{D_n}I_{inc,r} - \frac{1}{D_{inc}}I_{inc,r} - \mu_{exit}\, I_{inc,r}$$

$\frac{dI_{s,r}}{dt}$ = symptom onset – innate recovery – treatment – population exit

$$\frac{dI_{s,r}}{dt} = \frac{1}{D_{inc}}I_{inc,r} - \frac{1}{D_n}I_{s,r} - \frac{1}{D_{treat}}\, I_{s,r} - \mu_{exit}\, I_{s,r}$$

$\frac{dI_{s,r,treat,success}}{dt}$ = successful treatment – recovery upon treatment – population exit

$$\frac{dI_{s,r,treat,success}}{dt} = (1 - p_{tf})\frac{1}{D_{treat}}\,(1 - p_d)I_{s,r} - \frac{1}{D_r}I_{s,r,treat,success} - \mu_{exit}I_{s,r,treat,success}$$

$\frac{dI_{s,r,treat,fail}}{dt}$ = failed treatment – retreatment – innate recovery - population exit

$$\frac{dI_{s,r,treat,fail}}{dt} = \frac{p_{tf}}{D_{treat}}(1 - p_d)I_{s,r} - \frac{1}{D_{rt}}I_{s,r,treat,fail} - \frac{1}{D_n}I_{s,r,treat,fail} - \mu_{exit}I_{s,r,treat,fail}$$

$\frac{dI_{s,r,detect}}{dt}$= detection at first treatment + retreatment – recovery upon treatment – population exit

$$\frac{dI_{s,r,detect}}{dt} = \frac{1}{D_{treat}}\, p_d I_{s,r} + \frac{1}{D_{rt}}I_{s,r,treat,fail} - \frac{1}{D_r}I_{s,r,detect} - \mu_{exit}I_{s,r,detect}$$

Asymptomatic infection with resistant strain:

$\frac{dI_{a,r}}{dt}$= infection – innate recovery – treatment – population exit

$$\frac{dI_{a,r}}{dt} = (1 - p_{symptoms}) \frac{\boldsymbol{\beta I_r} S}{N} - \frac{1}{D_n} I_{a,r} - \frac{1}{D_{screen}} I_{a,r} - \mu_{exit} I_{a,r}$$

$\frac{dI_{a,r,treat,success}}{dt}$ = successful treatment – recovery upon treatment – population exit

$$\frac{dI_{a,r,treat,success}}{dt} = (1 - p_{tf})(1 - p_d) \frac{1}{D_{screen}} I_{a,r} - \frac{1}{D_r} I_{a,r,treat,success} - \mu_{exit} I_{a,r,treat,success}$$

$\frac{dI_{a,r,treat,fail}}{dt}$ = failed treatment – innate recovery - retreatment – population exit

$$\frac{dI_{a,r,treat,fail}}{dt} = p_{tf}(1 - p_d) \frac{1}{D_{screen}} I_{a,r} - \frac{1}{D_n} I_{a,r,treat,fail} - \left(p_{TOC} + \frac{1}{D_{screen}}\right) I_{a,r,treat,fail} - \mu_{exit} I_{a,r,treat,fail}$$

$\frac{dI_{a,r,detect}}{dt}$ = detection at first treatment + retreatment – recovery upon treatment – population exit

$$\frac{dI_{a,r,detect}}{dt} = p_d \frac{1}{D_{screen}} I_{a,r} + p_{TOC} I_{a,r,treat,fail} - \frac{1}{D_r} I_{a,r,detect} - \mu_{exit} I_{a,r,detect}$$

<u>Symptomatic infection with non-resistant strain:</u>

$\frac{dI_{inc,nr}}{dt}$ = - population exit + infection – innate recovery – symptom onset

$$\frac{dI_{inc,nr}}{dt} = p_{symptoms} \frac{\boldsymbol{\beta S I_{nr}}}{N} - \frac{1}{D_n} I_{inc,nr} - \frac{1}{D_{inc}} I_{inc,nr} - \mu_{exit} I_{inc,nr}$$

$\frac{dI_{s,nr}}{dt}$ = symptom onset – innate recovery – treatment – population exit

$$\frac{dI_{s,nr}}{dt} = \frac{1}{D_{inc}} I_{inc,nr} - \frac{1}{D_n} I_{s,nr} - \frac{1}{D_{treat}} I_{s,nr} - \mu_{exit} I_{s,nr}$$

<u>Asymptomatic infection with non-resistant strain:</u>

$\frac{dI_{a,nr}}{dt}$= infection – innate recovery – treatment – population exit

$$\frac{dI_{a,nr}}{dt} = (1 - p_{symptoms}) \frac{\boldsymbol{\beta I_{nr}} S}{N} - \frac{1}{D_n} I_{a,nr} - \frac{1}{D_{screen}} I_{a,nr} - \mu_{exit} I_{a,nr}$$

$\frac{dI_{nr,treat,success}}{dt}$ = treatment – recovery upon treatment – population exit

$$\frac{dI_{nr,treat,success}}{dt} = (1 - p_d) \frac{1}{D_{treat}} I_{s,nr} + (1 - p_d) \frac{1}{D_{screen}} I_{a,nr} - \frac{1}{D_r} I_{nr,treat,success} - \mu_{exit} I_{nr,treat,success}$$

$\frac{dI_{nr,detect}}{dt}$ = detection at first treatment – recovery upon treatment – population exit

$$\frac{dI_{nr,detect}}{dt} = p_d \frac{1}{D_{treat}} I_{s,nr} + p_d \frac{1}{D_{screen}} I_{a,nr} - \frac{1}{D_r} I_{nr,detect} - \mu_{exit} I_{nr,detect}$$

Where:

Mixing matrix

$\beta_{ij} = b_{ij} m^*_{i \to j}$

$\mathrm{m_{i \to j}} = \epsilon\, \mathrm{c_i N_i} \frac{\mathrm{c_j N_j}}{\sum_{\mathrm{k} \in \mathrm{K_a}} \mathrm{c_k N_k}}$ for $i, j$ in same activity group, and $\mathrm{m_{i \to j}} = (1 - \epsilon)\, \mathrm{c_i N_i} \frac{\mathrm{c_j N_j}}{\sum_{\mathrm{k} \in \mathrm{K_d}} \mathrm{c_k N_k}}$ for $i, j$ in different activity groups, where $c_i$ is the average number of partners for individuals in group $i$, $N_i$ is the population size of group $i$, $\epsilon$ is the assortative mixing parameter, and $b_{ij}$ is the per-partnership transmission probability given an infectious contact. Mixing is defined in this way for activity groups within MSM, between MSW and WSM, between MSMW and MSM, and between MSMW and WSM.

Mixing is adjusted to balance supply and demand for partnerships:
$m^*_{i \to j} = B^{\theta - 1} m_{i \to j}$
$m^*_{j \to i} = B^{\theta} m_{j \to i}$
where $\theta$ is a balancing parameter and $B = \frac{m_{i \to j}}{m_{j \to i}}$.

Notation

$\boldsymbol{I} = \boldsymbol{I_{inc}} + \boldsymbol{I_s} + \boldsymbol{I_a} + \boldsymbol{I_{treat,success}} + \boldsymbol{I_{treat,fail}}$
$\boldsymbol{I_{inc}} = I_{inc,r} + I_{inc,nr}$
$\boldsymbol{I_a} = I_{a,r} + I_{a,nr}$
$\boldsymbol{I_s} = I_{s,r} + I_{s,nr}$
$\boldsymbol{I_{treat,fail}} = I_{a,r,treat,fail} + I_{s,r,treat,fail}$
$\boldsymbol{I_{treat,success}} = I_{a,r,treat,success} + I_{a,nr,treat,success} + I_{s,r,treat,success} + I_{s,nr,treat,success}$

Compartments:

$S$ : Susceptible
$I$ : Infectious

Subscripts:

$r$: resistant strain
$nr$: non-resistant strain
$a$: asymptomatic
$inc$: incubation period
$s$: symptomatic
$treat, success$: successful treatment
$treat, fail$: failed treatment
$detect$: treatment of infection and antibiotic susceptibility testing of strain

#### Table S1. Fixed Parameters

| Parameter | Description | Group | Value | Source |
|---|---|---|---|---|
| $N$ | Population size | | 5,679,309 | MDPH |
| | Population proportion, by group | MSM, high activity | 0.025 * 0.15 | Assumptions, following (1–4) |
| | | MSM, low activity | 0.025 * 0.85 | |
| | | MSMW, high activity | 0.025 * 0.15 | |
| | | MSMW, low activity | 0.025 * 0.85 | |
| | | MSW, high activity | 0.475 * 0.15 | |
| | | MSW, low activity | 0.475 * 0.85 | |
| | | WSM, high activity | 0.475 * 0.15 | |
| | | WSM, low activity | 0.475 * 0.85 | |
| $\mu_{entry}, \mu_{exit}$ | Rates of population entry and exit, per person per day | | 1/(20*365) | (3,5) |
| | Initial gonorrhea prevalence | MSM (all) | 0.03 | Assumption, following (3) |
| | | MSM, high activity | 0.08 | Assumption |
| | | Men | 0.0126 | (6) |
| | | Women | 0.0161 | (6) |
| | | Women, high activity | 0.027 | (7,8) |
| $p_{TOC}$ | Rate of follow-up for test of cure after asymptomatic screening | Men | 0.22 / 10 | (9–11) |
| | | Women | 0.38 / 10 | (9) |
| $D_{rt}$ | Time until retreatment, in days, given treatment failure of symptomatic infection | Men | 7 | Assumption |
| | | Women | 7 | Assumption |
| $p_{tf}$ | Probability of treatment failure if resistant strain is treated with ceftriaxone | | 0.8 | Assumption |

Abbreviations: Men who have sex with men (MSM), men who have sex with men and women (MSMW), men who have sex with women (MSW), women who have sex with men (WSM).

#### Table S2. Fitted Parameters

| Parameter | Description | Group | Prior distribution | Prior mean | Reference | Posterior mean (75% CI) |
|---|---|---|---|---|---|---|
| $p_{symptoms}$ | Proportion of infections that are symptomatic | Men | Beta(4.3, 2.3) | 0.65 | MDPH | 0.40 (0.31-0.51) |
| | | Women | Beta(6.2, 5.3) | 0.54 | MDPH | 0.53 (0.45-0.64) |
| $c_{i,high}$ for $i \in \{MSM, MSMW, MSW, WSM\}$ | Average number of contacts (per year), high activity groups | MSM | Gamma(9.3, 3.3) | 30.5 | (12), DPH | 30.9 (23.3-38.7) |
| | | MSMW | Male: Gamma(5.2,4.4) Female: Gamma(3.8, 4.0) | Male: 30.5*0.75 Female: 20*0.75 | Assumption | Male: 28.6 (17.6-37.2) Female: 18.7 (11.6-22.7) |
| | | MSW | Gamma(6.7, 3.0) | 20 | (5,13), MDPH | 18.4 (11.8-21.9) |
| | | WSM | Gamma(6.7, 3.0) | 20 | (5,13), MDPH | 20.5 (16.8-25.7) |
| $p_{L,i}$ where $c_{i,low} = p_{L,i} c_{i,high}$ for $i \in \{MSM, MSMW, MSW, WSM\}$ | Average number of contacts (per) year in low activity group, as a proportion of high activity group | MSM | Uniform(0,1) | 0.5 | | 0.48 (0.25-0.68) |
| | | MSMW | Uniform(0,1) | 0.5 | | Male: 0.58 (0.50-0.73) Female: 0.48 (0.27-0.72) |
| | | MSW | Uniform(0,1) | 0.5 | | 0.41 (0.14-0.66) |
| | | WSM | Uniform(0,1) | 0.5 | | 0.40 (0.10-0.64) |
| $\epsilon$ | Assortative mixing parameter | | Beta(2,2) | 0.5 | (14) | 0.49 (0.30-0.69) |
| $b_{ij}$ | Per-partnership transmission probability given infectious contact between groups $i, j$ | MSM, low-low | Beta(13.7, 9.5) | 0.59 | (12,15) | 0.61 (0.55-0.69) |
| | | MSM, high-high | Beta(12.0,12.0) | 0.5 | (3,12,15) | 0.53 (0.47-0.59) |
| | | Male-to-female, low-low | Beta(12.0,3.0) | 0.8 | (12,13) | 0.80 (0.74-0.89) |
| | | Male-to-female, high-high | Beta(13.8, 5.4) | 0.72 | (12) | 0.72 (0.64-0.79) |
| | | Female-to-male, low-low | Beta(12.0,3.0) | 0.8 | (12,13) | 0.77 (0.73-0.82) |
| | | Female-to-male, high-high | Beta(13.8, 5.4) | 0.72 | (12) | 0.73 (0.68-0.80) |
| $D_{treat}$ | Duration from start of | Men | Gamma(6.7, 0.8) | 5.3 | MDPH | 5.6 (4.1-6.4) |

| | | | | | | |
|---|---|---|---|---|---|---|
| | symptoms to treatment, in days | Women | Gamma(13.6, 0.6) | 8.4 | MDPH | 8.5 (7.1-10.0) |
| $D_r$ | Duration from time of treatment to recovery | Men | Gamma(3.0, 1.0) | 3 | (16) | 3.2 (2.2-4.0) |
| | | Women | Gamma(3.0, 1.0) | 3 | (16) | 3.0 (1.8-3.6) |
| $D_n$ | Duration from infection to natural clearance | Men | Gamma(484.0, 0.1) | 44 | (17–19) | 44.1 (42.7-45.3) |
| | | Women | Gamma(1936.0, 0.05) | 88 | (17–19) | 87.7 (86.5-87.7) |
| $D_{inc}$ | Duration of incubation period | Men | Gamma(22.4, 0.3) | 6.7 | (17,18,20) | 6.8 (6.1-7.6) |
| | | Women | Gamma(36.0, 0.3) | 12 | (17,18,21) | 12.2 (10.8-13.0) |
| $F_{screen,i}$ , where $D_{screen,high,i} = \frac{365}{F_{screen,i}}$ for $i \in \{MSM, MSMW, MSW, WSM\}$ | Average asymptomatic screening frequency (per year) | MSM, high activity | Uniform(1,4) | 2.5 | (22) | 2.5 (1.7 - 3.2) |
| | | MSMW, high activity | Uniform(1,4) | 2.5 | (22) | 2.3 (1.8 – 2.8) |
| | | MSW, high activity | Uniform(0,1) | 0.5 | (22) | 0.57 (0.37-0.86) |
| | | WSM, high activity | Uniform(0,1) | 0.5 | (22) | 0.44 (0.23-0.65) |
| $p_{screen,low,i}$ where $D_{screen,low,i} = \frac{365}{p_{screen,low,i} F_{screen,i}}$ for $i \in \{MSM, MSMW, MSW, WSM\}$ | Average asymptomatic screening frequency, (per year) | MSM, low activity | Uniform(0,1) * $F_{screen,MSM}$ | 0.5 * $F_{screen,MSM}$ | | 1.4 (0.6-2.0) |
| | | MSMW, low activity | Uniform(0,1) * $F_{screen,MSMW}$ | 0.5 * $F_{screen,MSMW}$ | | 1.1 (0.6-1.7) |
| | | MSW, low activity | Uniform(0,1) * $F_{screen,MSW}$ | 0.5 * $F_{screen,MSW}$ | | 0.3 (0.1-0.4) |
| | | WSM, low activity | Uniform(0,1) * $F_{screen,WSM}$ | 0.5 * $F_{screen,WSM}$ | | 0.2 (0.1-0.3) |
| $p_{detect}$ | Antibiotic susceptibility testing probability (per positive sample) | Men | Uniform(0,0.3) | 0.15 | Assumption, MDPH | 0.05 (0.03-0.06) |
| | | Women | Uniform(0,0.3) | 0.15 | Assumption, MDPH | 0.03 (0.01-0.04) |

Abbreviations: Men who have sex with men (MSM), men who have sex with men and women (MSMW), men who have sex with women (MSW), women who have sex with men (WSM).

#### Table S3. Calibration targets

| Target | Group | Accepted range | Source |
|---|---|---|---|
| Annual observed incidence, per 100,000 population | Men | $(0.75 * 181; 1.25 * 181\ )$ | MDPH |
| | Women | $(0.75 * 80; 1.25 * 80)$ | MDPH |
| Prevalence, per population | MSM | $(0.02; 0.06)$ | (3) |
| | MSW | $(0.003; 0.02)$ | (12,23–25) |
| | WSM | $(0.003; 0.02)$ | (12,23–25) |

Abbreviations: Men who have sex with men (MSM), men who have sex with men and women (MSMW), men who have sex with women (MSW), women who have sex with men (WSM).

#### Supplemental Results

**Figure S2: Disease burden and likelihood of strain elimination for increasing days without detection of resistant cases, following the detection of 2 resistant cases**
(A) Estimate of the number of undetected infections with the resistant strain after increasing days without newly reported resistant cases (mean and 95% uncertainty interval).
(B) Proportion of simulations without new cases for increasing days without newly reported resistant cases (mean and 95% uncertainty interval). Dashed lines highlight values at 0, 60, and 180 days without detection.

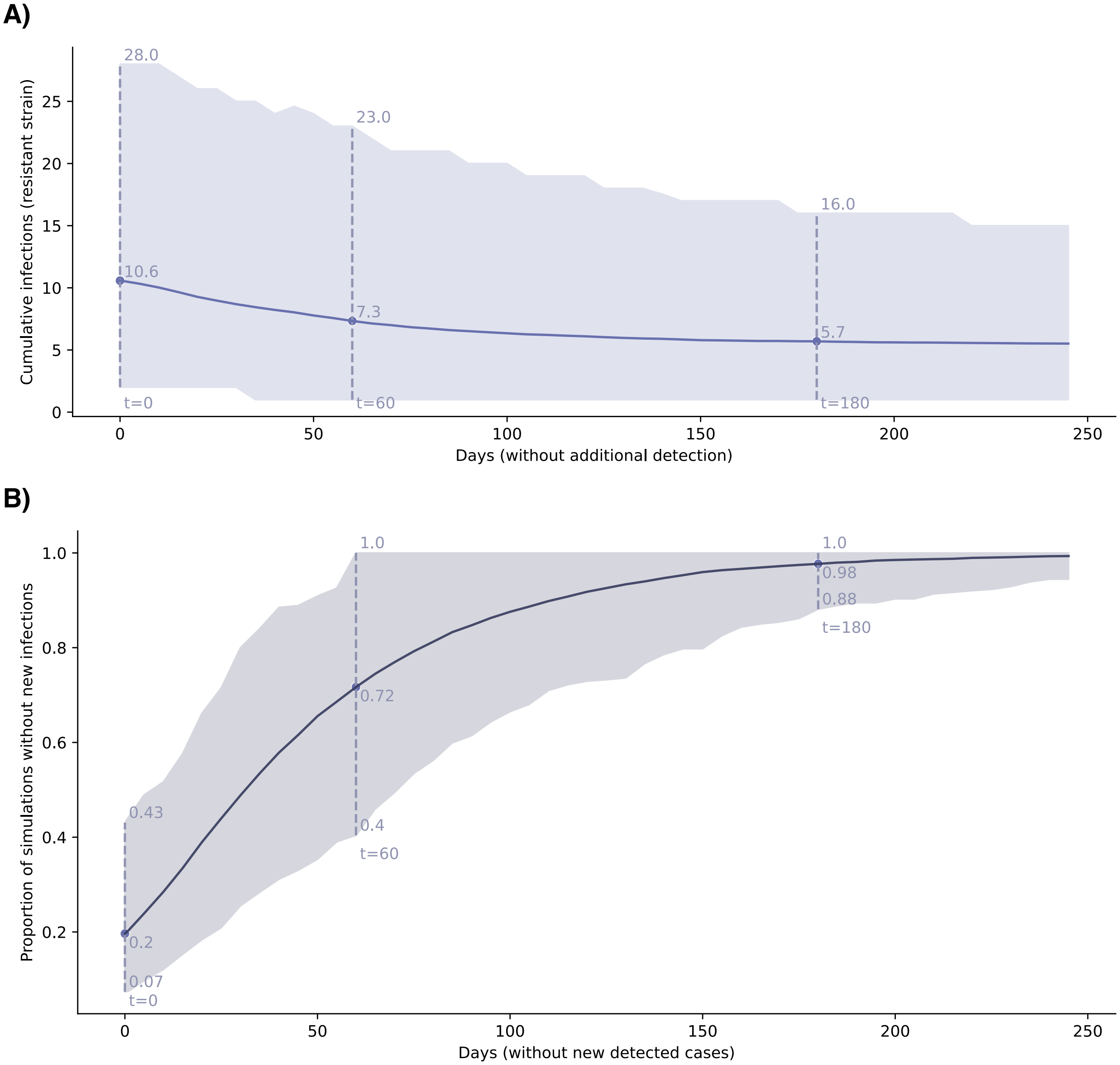

**Figure S3. Confidence in elimination for varying levels of detected cases.** Proportion of simulations without new cases for increasing days without newly reported resistant cases after initially detecting 1, 5, and 10 resistant cases (mean and 95% uncertainty interval).

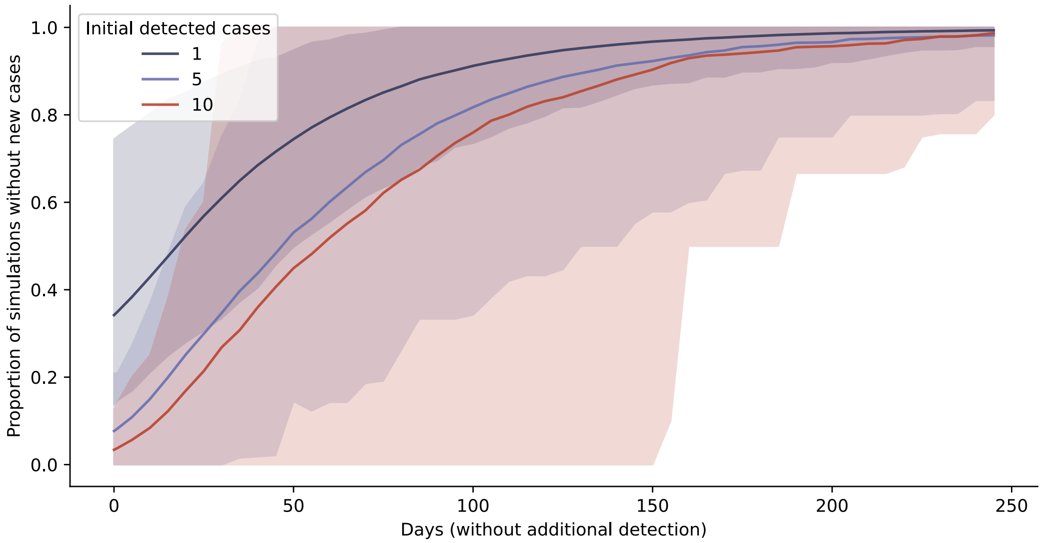


**Figure S4. Undetected disease burden for varying treatment failure rates.**
Undetected non-susceptible infections upon detection of one non-susceptible case and increasing days without newly detected cases (mean and 95% uncertainty interval), for varying treatment failure rates.

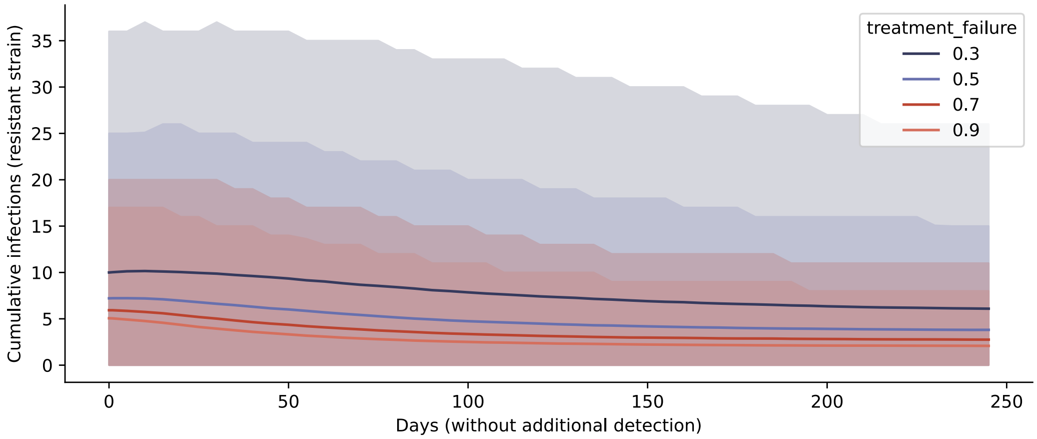


**Figure S5. Undetected disease burden for varying surveillance intensities.**
Undetected non-susceptible infections upon detection of one non-susceptible case and increasing days without newly detected cases (mean and 95% uncertainty interval), for (A) varying antibiotic susceptibility testing rates and (B) varying asymptomatic screening intensities.

**A) Varying antibiotic susceptibility testing rates**

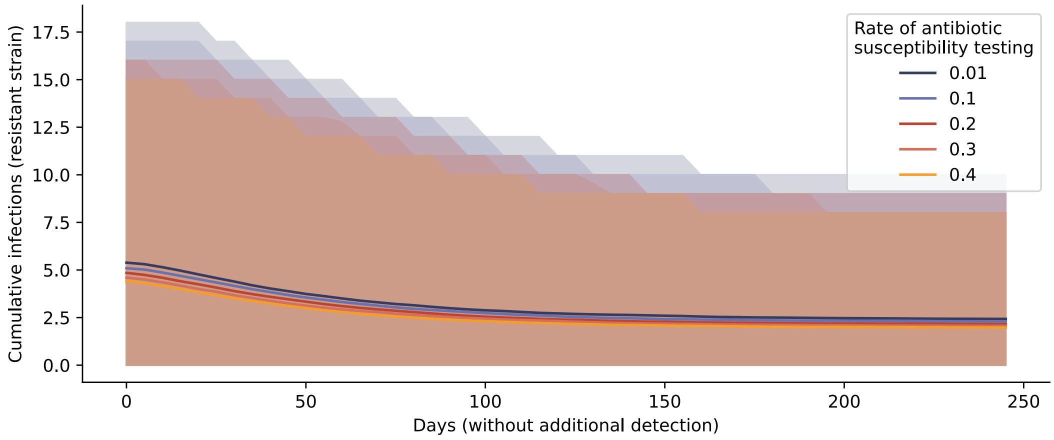


#### B) Varying asymptomatic screening intensities

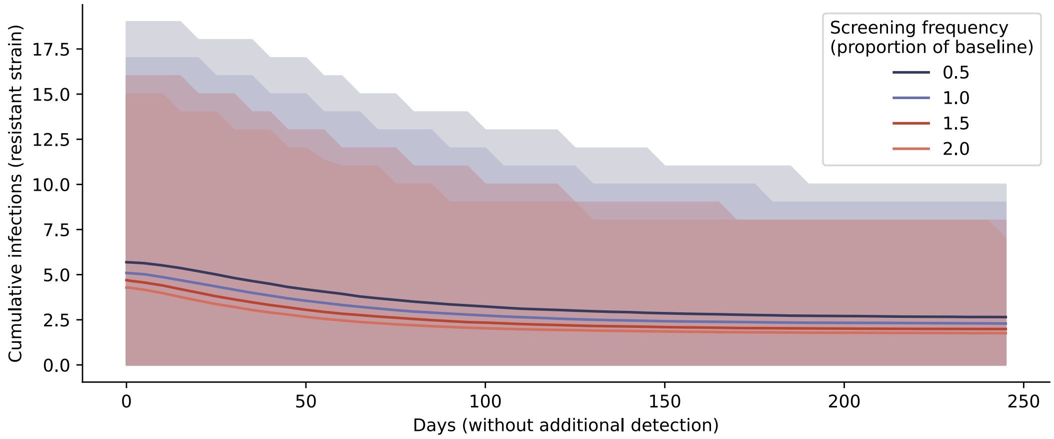
